# Seroprevalence of antibodies against *Chlamydia* trachomatis and enteropathogens and distance to the nearest water source among young children in the Amhara Region of Ethiopia

**DOI:** 10.1101/2020.04.16.20060996

**Authors:** Kristen Aiemjoy, Solomon Aragie, Dionna M. Wittberg, Zerihun Tadesse, E. Kelly Callahan, Sarah Gwyn, Diana Martin, Jeremy D. Keenan, Benjamin F. Arnold

## Abstract

**Background:** The transmission of trachoma, caused by repeat infections with *Chlamydia trachomatis*, and many enteropathogens are linked to water quantity. We hypothesized that children living further from a water source would have higher exposure to *C. trachomatis* and enteric pathogens as determined by antibody responses.

**Methods:** We used a multiplex bead assay to measure IgG antibody responses to *C. trachomatis, Giardia intestinalis, Cryptosporidium parvum, Entamoeba histolytica, Salmonella enterica, Campylobacter jejuni*, enterotoxigenic *Escherichia coli* (ETEC) and *Vibrio cholerae* in eluted dried blood spots collected from 2267 children ages 1–9 years in 40 communities in rural Ethiopia in 2016. Linear distance from the child’s house to the nearest water source was calculated. We derived seroprevalence cutoffs using external negative control populations, if available, or by fitting finite mixture models. We used targeted maximum likelihood estimation to estimate differences in seroprevalence according to distance to the nearest water source.

**Results:** Seroprevalence among 1–9-year-olds was 43% for *C. trachomatis*, 28% for *S. enterica*, 70% for *E. histolytica*, 54% for *G. intestinalis*, 96% for *C. jejuni*, 76% for ETEC and 94% for *C. parvum*. Seroprevalence increased with age for all pathogens. Median distance to the nearest water source was 473 meters (IQR 268, 719). Children living furthest from a water source had a 12% (95% CI: 2.6, 21.6) higher seroprevalence of *S. enterica* and a 12.7% (95% CI: 2.9, 22.6) higher seroprevalence of *G. intestinalis* compared to children living nearest.

**Conclusion:** Seroprevalence for *C. trachomatis* and enteropathogens was high, with marked increases for most enteropathogens in the first two years of life. Children living further from a water source had higher seroprevalence of *S. enterica and G. intestinalis* indicating that improving access to water in the Ethiopia’s Amhara region may reduce exposure to these enteropathogens in young children.

**AUTHOR SUMMARY:** Trachoma, and infection of the eye caused by the bacteria *Chlamydia trachomatis*, and many diarrhea-causing infections are associated with access to water for washing hands and faces. Measuring these different pathogens in a population is challenging and rarely are multiple infections measured at the same time. Here, we used an integrated approach to simultaneously measure antibody responses to *C. trachomatis, Giardia intestinalis, Cryptosporidium parvum, Entamoeba histolytica, Salmonella enterica, Campylobacter jejuni*, enterotoxigenic *Escherichia coli* (ETEC) and *Vibrio cholerae* among young children residing in rural Ethiopia. We found that the seroprevalence of all pathogens increased with age and that seropositivity to more than one pathogen was common. Children living further from a water source were more likely to be exposed to *S. enterica and G. intestinalis*. Integrated sero-surveillance is a promising avenue to explore the complexities of multi-pathogen exposure as well as to investigate the relationship water, sanitation and hygiene related exposures disease transmission.

## BACKGROUND

Diarrhea and trachoma typically afflict the world’s poorest populations and are major causes of preventable morbidity [1,2]. Diarrhea, caused by parasitic, viral and bacterial infections, and trachoma, caused by repeated *Chlamydia trachomatis* infections of the eye, share water and hygiene related transmission pathways. Increased access to water for food preparation and washing of hands, faces, and clothing is hypothesized to reduce transmission of both infectious diarrhea and *C. trachomatis* [3–6]. In regions where water must be carried from the source to the household, distance to the nearest water source will likely influence the quantity of water a household uses [7].

Antibody responses may be an informative and efficient approach to simultaneously measure enteropathogen and *C. trachomatis* exposure [8–10]. Unlike pathogen detection from stool samples or conjunctival swabs, antibody response integrates information over time, offering a longer window to identify exposed individuals. [9]. This advantage is especially desirable for studies with infrequent monitoring and data collection visits. Antibody response enumerates symptomatic, asymptomatic and past infections, revealing a more complete picture of transmission [9]. With recent advances in microsphere-based multiplex immunoassays, antibodies against multiple antigens can be detected simultaneously from a single blood spot [11]. This technology has a unique advantage that it can be used to simultaneously monitor for dozens of markers of pathogen transmission, potentially revealing vulnerable populations and/or individuals who experience the pervasive burdens of multiple-pathogen exposure.

In this study we evaluated IgG antibody responses to a panel of antigens from viral, bacterial, and protozoan enteropathogens and *C. trachomatis* antigens among a population-based cohort of children aged 0 to 9 years in rural Ethiopia. Our objectives were to describe age-dependent seroprevalence and co-prevalence of the pathogens and to evaluate if seroprevalence varied according to distance to nearest water source.

## METHODS

### Study design overview

We conducted a cross-sectional study evaluating antibody responses in children at the baseline visit of a cluster-randomized trial of a water, sanitation and hygiene (WASH) intervention in 40 communities in the Amhara region of Ethiopia. We used a multiplex bead assay to simultaneously measure IgG antibodies to antigens from *Chlamydia trachomatis* (Pgp3, CT694), *Giardia intestinalis (*VSP3, VSP5*), Cryptosporidium parvum* (Cp17, Cp23), *Entamoeba histolytica* (LecA), *Salmonella enterica* (LPS Groups B and D), *Campylobacter jejuni* (p18, p39), enterotoxigenic *Escherichia coli* (ETEC heat labile toxin β subunit) and *Vibrio cholerae* (CtxB) from blood spots collected during the baseline study visit.

### Study population

Sanitation, Water, and Instruction in Face-washing for Trachoma (SWIFT), is an ongoing NIH-funded cluster-randomized trial designed to determine the effectiveness of a comprehensive WASH package for ocular *C. trachomatis* infection (ClinicalTrials.gov: NCT02754583) in three *woredas* (districts) of the WagHemra zone of Amhara, Ethiopia. This cross-sectional analysis was carried out during the baseline study visit for SWIFT, from January to April 2016.

Study staff performed a door-to-door census in December 2015, approximately one month before the baseline examination visit. Census workers recorded the name, sex, and age of each household member and the GPS coordinates of the house. From this census, we drew a random sample of 30 children aged 0 to 5 years and 30 children aged 6 to 9 years in each cluster for inclusion in the study. The sample size was calculated for the primary outcome of the trial (ocular *C. trachomatis* infection).

### Measurements

#### Dried blood spots

A few days before each study visit a volunteer was sent out into the community to mobilize sampled children and their accompanying caregivers to attend the examination visit, with information on the time and location of the event. A trained laboratory technician lanced the index finger of each child and collected 5 blood spots onto a TropBioTM filter paper (Cellabs Pty Ltd., Brookvale, New South Wales, Australia) calibrated to hold 10 µL of blood per spot. The filter paper was labeled with a random number identification sticker, air-dried for at least one hour and then individually packaged in plastic re-sealable bags. The individual bags from each cluster were placed in large, re-sealable bags with desiccant. The samples were stored at −20°C until all sample collection for the entire study visit was completed and then shipped at ambient temperature to the Centers for Disease Control and Prevention (CDC) in Atlanta, GA, where they were stored at −20°C until testing.

#### Distance to water

At the time of the census, census workers asked community leaders to list all sources of water used in the community. The census workers then visited each water source, recorded the GPS coordinates and described the type of water source. Linear distance to the nearest water source was calculated from the household using GPS coordinates. We hypothesized that the quantity of water available to the household would have a larger effect on *C. trachomatis* and enteropathogen transmission compared to water quality, and thus used distance to the nearest water source (improved or unimproved) for the analysis.

#### Covariates

In a random one-third of households, study field workers performed a household survey evaluating socioeconomic status, access to water, number/type of animals in household, hygiene behaviors, and sanitation infrastructure.

### Laboratory methods

We measured IgG responses against *C. trachomatis* and enteropathogen antigens using a multiplex SeroMAPTM microsphere-based immunoassay on the Luminex xMAP platform (Luminex Corp, Austin, TX) for the following antigens: *G. intestinalis* variant-specific surface protein AS8/GST fusion (VSP3) and 42e/GST fusion (VSP5) [12–14]; *C. jejuni antigen* p39 and p18 [15]; *Enterotoxigenic Escherichia coli* (ETEC heat labile toxin B subunit); *C. parvum* 17-kDa protein/GST fusion (Cp17) and 23-kDa protein/GST fusion (Cp23)[16]; *Salmonella spp. (*LPS Groups B and D) [9]; *V. cholerae* toxin B subunit (CtxB); *E. histolytica* Gal/GalNAc lectin heavy chain subunit (LecA) [17,18]; and *C. trachomatis* Pgp3 & CT694 [19]. *<u>Serum elution:</u>* The dried blood spots were brought to room temperature and submerged in 1600 μL of elution buffer for a minimum of 18 hours at 4°C [20]. *<u>Multiplex bead assay:</u> Each* 96-well plate included a buffer-only blank, one negative control, and two positive controls. The background from the buffer-only blank is subtracted from the result for each antigen, and values are reported as an average median fluorescence intensity with background subtracted (MFI-bg) [12,17].

### Statistical Analysis

For pathogens with two antigens (*C. trachomatis, G. intestinalis, C. parvum, C. jejuni* and *S. enterica*), children positive to either antigen were considered exposed.

Positivity cutoffs were defined using external control populations when available. For *C. trachomatis* Pgp3 & CT694 cutoffs were derived using ROC curves [20], for *C. parvum* Cp17 & Cp23 cutoffs were derived using a standard curve and for *G. intestinalis* VSP-3 & VSP-5 and *E. histolytica* LecA cutoffs were derived using the mean plus 3 standard deviations above a negative control panel [21]. For the remaining antigens we used finite mixture models to fit Gaussian distributions for the log^10^ transformed MFI-bg values [10,22] and determined the seropositivity cutoffs using the mean plus three standard deviations of the first component. When estimating seropositivity cutoffs using mixture models, we restricted the population to children age 0 to 2 years at the exam date to ensure a sufficient number of unexposed children (Supplemental Figure 1) [10].

For the descriptive seroprevalence analyses we included all sampled children (aged 0 to 9 years old). In the analysis of antibody response and distance to water source we restricted the age range to 0 to 3 years for most enteropathogens because there was almost no outcome heterogeneity above age 3, consistent with other enteropathogen serology in cohorts from low-resource settings [10]. For pathogens with presumed lower transmission based on more slowly rising age-dependent seroprevalence (*C. trachomatis* and *S. enterica*) we used the full age range (0 to 9 years). All age ranges were pre-specified.

We estimated age-dependent seroprevalence using two complementary approaches. First, we used a stacked ensemble machine learning algorithm called “super learner” that combines predictions from multiple algorithms to ensure the best estimate of the age-dependent seroprevalence [23]. We included the following algorithms in the library: the simple mean, generalized linear models (GLMs), locally weighted regression (loess), generalized additive models with natural splines, and random forest. The super learner algorithm weights each member of the library so that the combined prediction from the ensemble minimizes the cross-validated mean squared error. Ensemble fits of age-antibody curves do not converge at the standard *n*^1/2^ rate so pointwise confidence intervals are difficult to estimate [24]. We therefore also estimated the age-dependent antibody curves using a cubic spline for age within a generalized additive model (GAM) [25]. We estimated approximate simultaneous confidence intervals around the curves using a parametric bootstrap of the variance-covariance matrix of the fitted model parameters [26,27].

To estimate differences in seroprevalence according to distance to the nearest water source, we used doubly robust targeted maximum likelihood estimation (TMLE) with influence-curve based standard errors that treated clusters as the independent unit of analysis [9,28–30]. We calculated prevalence differences comparing the prevalence in the furthest (fourth) quartile of distance to the nearest water source to the prevalence in the nearest (first) quartile. This comparison was prespecified. When comparing children living in the two quartiles we were restricted to roughly half of the sample size. We included the same algorithms as above in the TMLE super learner library to adjust for age and other potential confounders. Among the 33% of children whose household was randomly selected for inclusion in the household survey, we adjusted for socio-economic status (SES) using an indicator variable calculated using a principal component analysis. We also compared differences in quantitative antibody response according to distance quartile using the same approach.

The analysis plan was pre-specified and is available through the open science framework (<u>osf.io/2r7tj</u>). All analyses were done in *R* (version 3.4.2).

### Ethics statement

Ethical approval for this study was granted by the National Research Ethics Review Committee of the Ethiopian Ministry of Science and Technology, the Ethiopian Food, Medicine, and Health Care Administration and Control Authority, and institutional review boards at the University of California, San Francisco and Emory University. CDC staff did not have contact with study participants or access to personal identifying information and were therefore determined to be non-engaged. Community leaders provided verbal consent before enrollment of the community in the trial. Verbal consent was obtained from each participant or their guardian prior to participation.

## RESULTS

We collected dried blood spots from 2267 children residing in 40 communities between January and March of 2016. The median age was 5 years (IQR 3–7); 51.3% (1169/2267) of children were female. The median distance to the nearest water source was 448 meters (IQR 268–719). The majority of children, 56.9% (1291/2267), lived in households whose nearest water source was unprotected. Household demographic information was available for 755 children. In this subset, 8.7% (66/755) of children lived in households with electricity, 10.1% (76/755) lived in households with a radio, 0% (0/761) lived in households with a mobile phone, 84.4% (637/755) lived in households that owned animals (Table 1).

**Table 1:** Population characteristics in overall population and subset with household survey

|  | Overall sample | Subset with household survey |
| --- | --- | --- |
| N() Children | 2267 | 755 |
| N() Communities | 40 | 40 |
| Median age (IQR) | 5 (3-7) | 5 (3-7) |
| Female | 1169 (51.6%) | 377 (49.9%) |
| Median distance (meters) to nearest water source (IQR) | 472.83 (268-719.2) | 482.24 (267.59-737.22) |
| **Nearest water source** |  |  |
| Open Water | 1195 (52.7%) | 426 (56.4%) |
| Unprotected Well | 76 (3.4%) | 26 (3.4%) |
| Protected Spring | 548 (24.2%) | 173 (22.9%) |
| Protected Well | 448 (19.8%) | 130 (17.2%) |
| **Household Characteristics** |  |  |
| Household has Electricity | • | 66 (8.7% ) |
| Household has Radio | • | 76 (10.1% ) |
| Household owns Animals | • | 637 (84.4% ) |
| Household has mobile phone | • | 0 (0%) |

The seroprevalence among 0–9 year-olds was 43.1% (95% CI: 38, 48.4) for *C. trachomatis*, 27.5% (95% CI: 23.6, 31.6) for *S. enterica*, 70.3% (95% CI:67.7, 72.8) for *E. histolytica*, 53.9% (95% CI: 51.8, 56.0) for *G. intestinalis*, 95.6% (95% CI: 94.4, 96.5) for *C. jejuni*, 76.3% (95% CI: 74.1, 78.4) for ETEC and 94% (95% CI: 92.8, 94.9) for *C. parvum*. Seroprevalence increased with age with marked differences across pathogens (Figure 1). For ETEC, *E. histolytica, C. parvum, C. jejuni* and *G. intestinalis*, over 70% of children were positive at age 2 years. The age-dependent seroprevalence slopes were less steep for both *C. trachomatis* and *S. enterica*; by age 9 over 60% of children were seropositive for *C. trachomatis* and over 40% of children were seropositive for *S. enterica*. Seropositivity for more than 1 pathogen was common (Figure 2). At age 2 years, the median number of pathogens to which a child was seropositive was 4 (IQR 3–5), increasing to 5 (IQR 4–6) by age 4 years.

**Figure 1:**
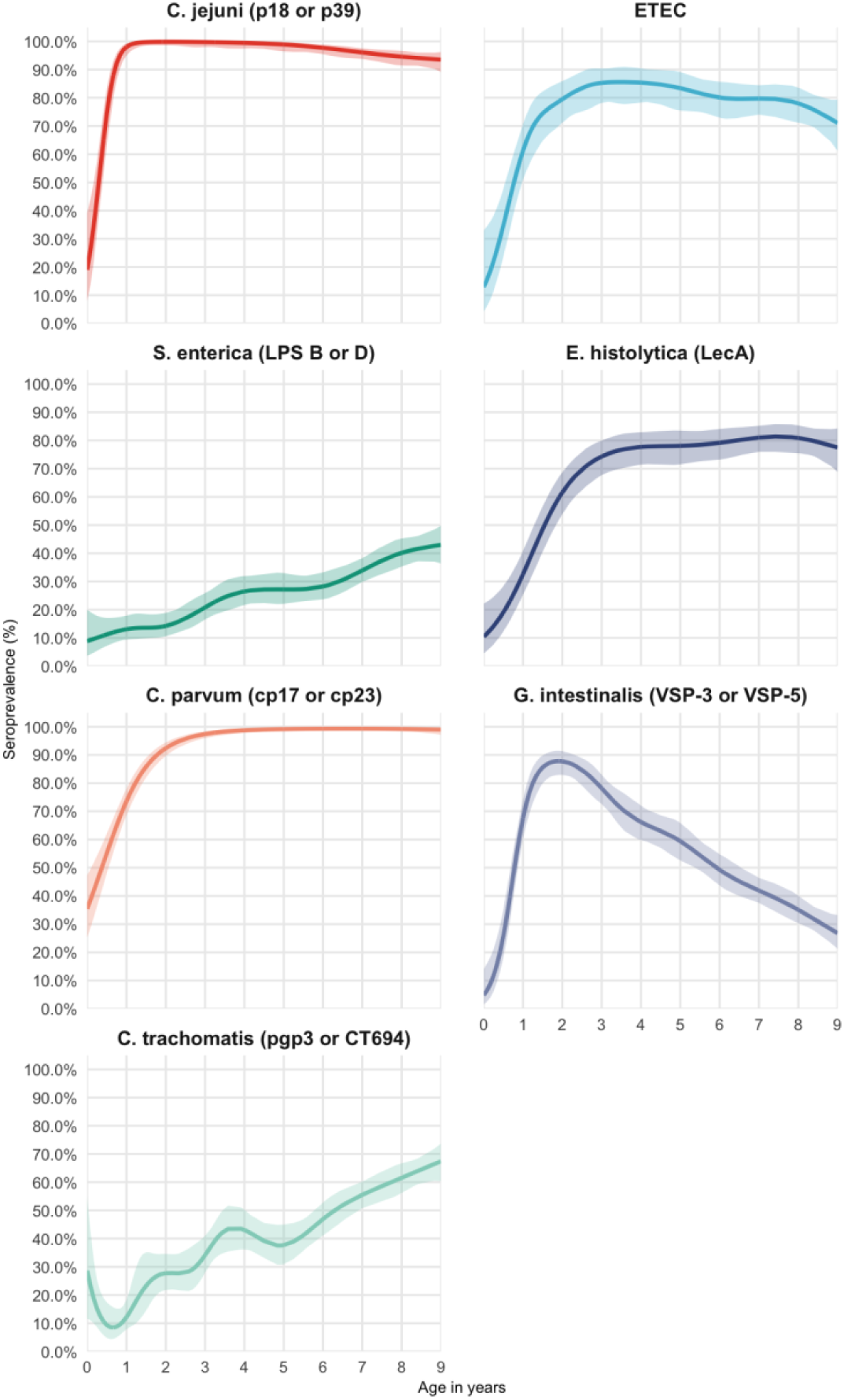
Age-dependent seroprevalence of trachoma and enteropathogens in the Amhara region of Ethiopia. Age-dependent seroprevalence curves were fitted using generalized additive models (GAM) with a cubic spline for age. Seropositivity cutoffs were derived using ROC curves, if available, or by fitting finite mixture models (Supplemental Figure 1). Seropositivity cutoffs could not be estimated for V. cholerae in this study, so seroprevalence curves are not shown. For pathogens with more than one antigen, positivity to antigen was considered positive. IgG response measured in multiplex using median fluorescence units minus background (MFI-bg) on the Luminex platform on 2267 blood samples from 2267 children.

**Figure 2:**
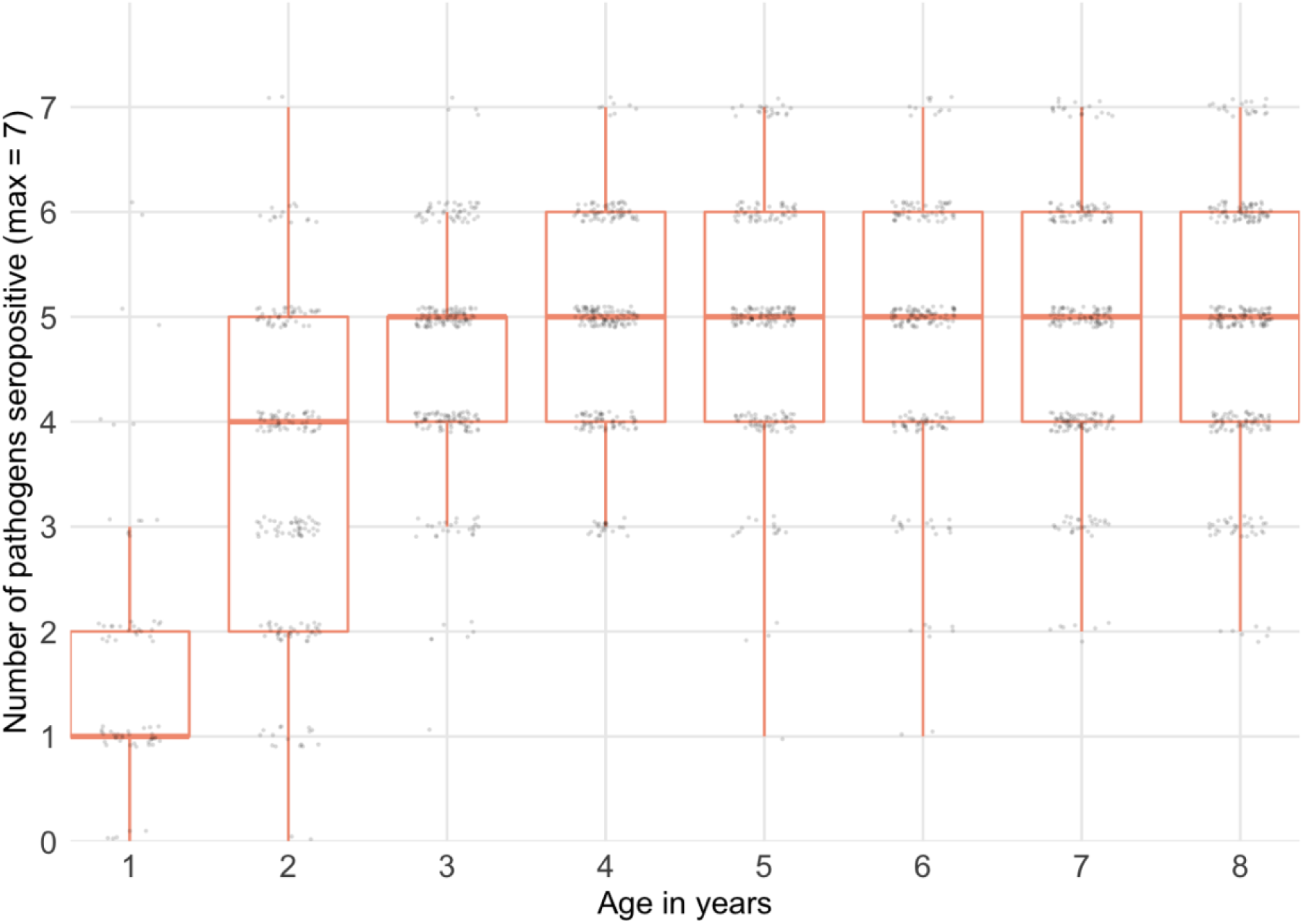
Seropositivity for more than 1 pathogen by age. Boxplot depicts median, upper and lower quartiles. Seropositivity cutoffs were derived using ROC curves, if available, or by fitting finite mixture models (Supplemental Figure 1). IgG response measured in multiplex using median fluorescence intensity minus background (MFI-bg) on the Luminex platform on 2267 blood samples from 2267 children.

There was no indication for trend in community-level seroprevalence by community-level median distance to the nearest water source; however, there was considerable variability on community-level seroprevalence for some pathogens (*C. trachomatis, G. intestinalis, E. histolytica and S. enterica* (Figure 3)). The between-community variance in seroprevalence was highest for *C. trachomatis* (SD .20) and *S. enterica* (SD 0.13). More community-level heterogeneity was apparent among young children (under 3) compared with older children, the exceptions being *C. parvum* and *C. jejuni* which both had very high seroprevalence even among young children. Correlation between community-level seroprevalence illustrated variation in co-occurrence (Supplemental Figure 2). There was indication for correlation between *C. trachomatis* and *E. histolytica*, ETEC, *C. jejuni* and *S. enterica* (Pearson correlation > 0.3).

**Figure 3:**
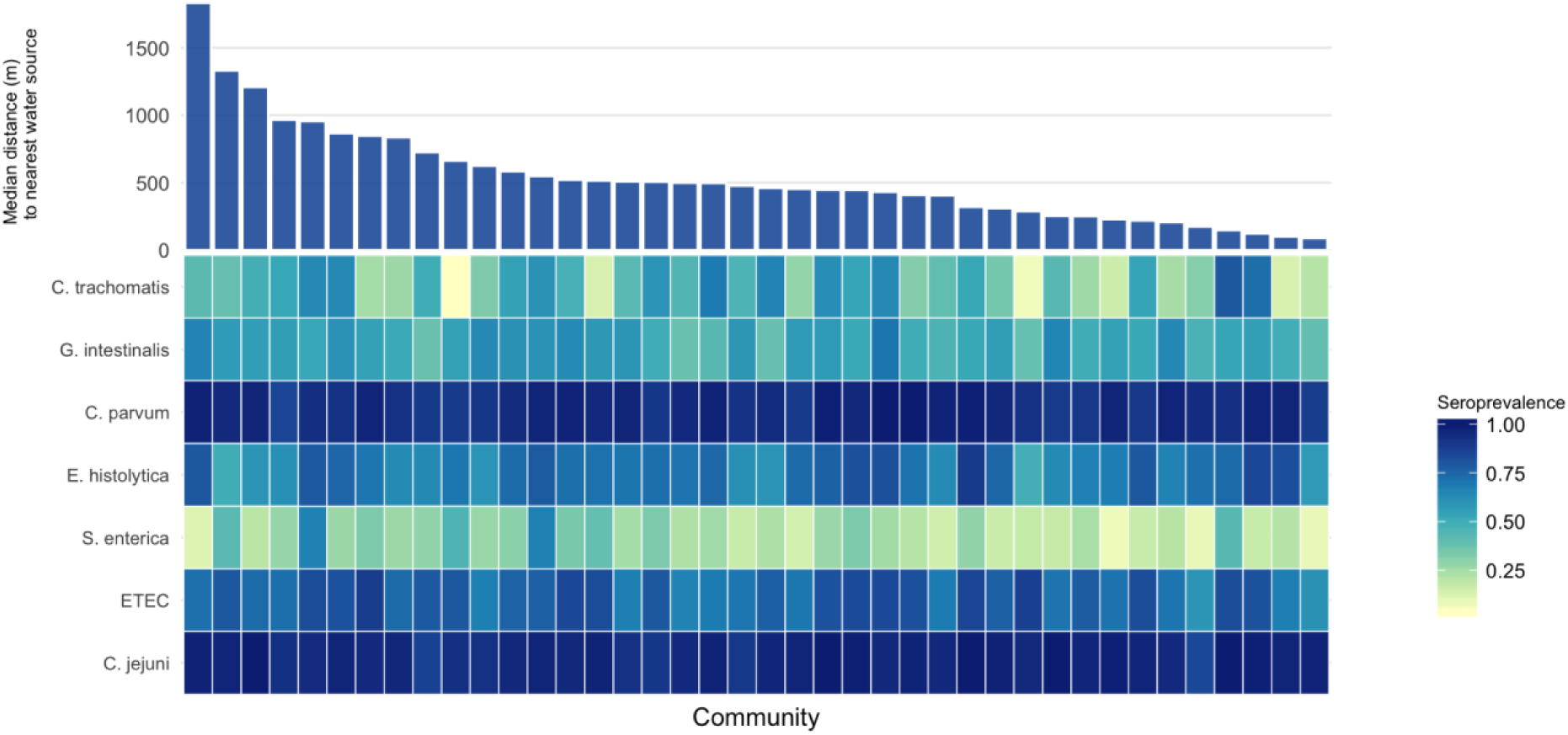
Variation in seroprevalence by community and distance to the nearest water source. Heatmap of community-level seroprevalence, darker colors indicate higher seroprevalence. Communities are sorted by median distance to the nearest water source, from furthest to nearest. Seropositivity cutoffs were derived using receiver operating characteristic (ROC) curves, if available, or by fitting finite mixture models (Supplemental Figure 1). For pathogens with more than one antigen, positivity to antigen was considered positive. IgG response measured in multiplex using median fluorescence intensity minus background (MFI-bg) on the Luminex platform on 2267 blood samples from 2267 children aged 0 to 9 years.

Children in the quartile living farthest from any water source had a 12% (95% CI: 2.6, 21.4) higher seroprevalence of *S. enterica* and a 12.7% (95% CI: 2.9, 22.6) higher seroprevalence of *G. intestinalis* compared to children living in the nearest quartile. The seroprevalence of ETEC and *C. trachomatis* were higher among children living in the furthest quartile of distance to the nearest water source, however the differences were not statistically significant (Table 2). Quantitative antibody levels demonstrated the same pattern for *S. enterica*, with antibody levels for *S. enterica* LPS group D 0.32 (95% CI: 0.13, 0.52) log^10^ MFI-bg units higher among children living in the furthest quartile from water compared to children living in the nearest quartile (p=0.001) (Supplemental Table 1). Quantitative antibody levels for ETEC and *G. intestinalis* were slightly higher among children living in the furthest quartile, but the differences were not statistically significant.

**Table 2:**
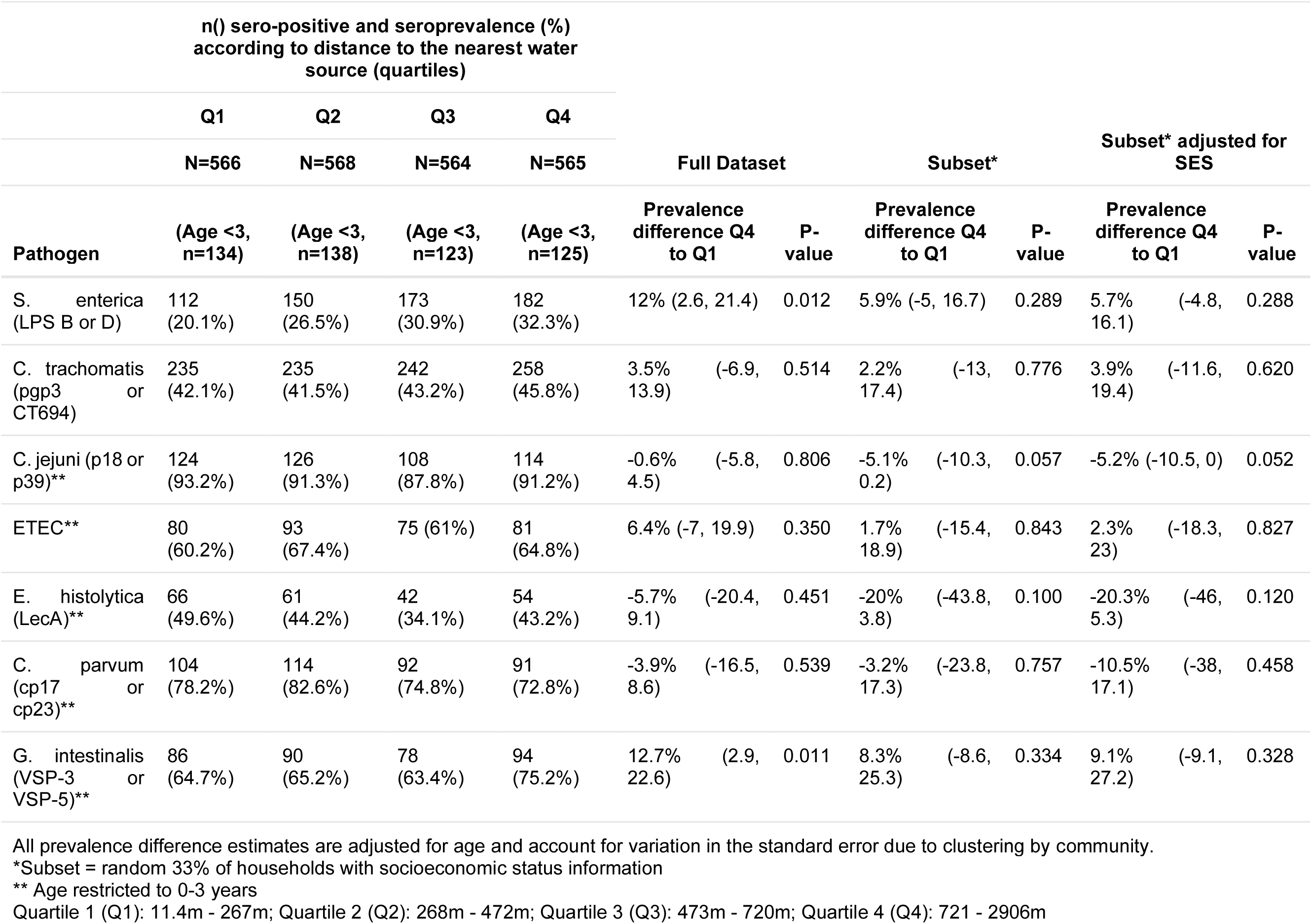
Seroprevalence according to distance to the nearest water source

In the subset of children with household-level data, point estimates were similar but there was no longer a statistically-significant association between distance to the nearest water source and seroprevalence for *S. enterica*, ETEC or *G. intestinalis* in the unadjusted or SES-adjusted analysis largely due to the smaller sample size and wider confidence intervals (Table 2).

## DISCUSSION

This study found high exposure to *C. trachomatis* and enteric pathogens among children residing in rural areas of the Amhara region of Ethiopia. Seroprevalence was age-dependent, with over 70% of children seropositive for ETEC, *E. histolytica, C. parvum, C. jejuni* and *G. intestinalis* at age two years. Age-dependent seroprevalence rose more slowly for *S. enterica* and *C. trachomatis*, suggesting lower transmission compared with the other enteropathogens. Still, at age 9 years, over 60% of children were seropositive for *C. trachomatis* and over 40% of children were seropositive for *S. enterica*. Children living farther from a water source had higher seroprevalence of *S. enterica* and *G. intestinalis*.

Use of a multiplexed immunoassay allowed us to expediently identify that seropositivity to more than one pathogen was common in the Amhara region of Ethiopia and that, by age three, most children were seropositive for five of the seven pathogens under investigation. Similarly, we were able to identify notable correlation in seroprevalence between some pathogens (for example, *C. parvum* and *E. histolytica)* at the community level. The seroprevalence of *G. intestinalis* and *E. histolytica* in this study was substantially higher than the prevalence reported in studies using microscopy in the region. In one recent study of protozoan prevalence in the Amhara region, the single-stool prevalence of *Entamoeba spp. (histolytica and dispar)* by microscopy among three year old children was 7.1% [31]. However, differences between seroprevalence and prevalence by microscopy are expected given that IgG response integrates information over time and microscopy measures active presence and shedding. The seroprevalence of trachoma identified in this study is consistent with the high burden of trachoma documented in the Amhara region [32].

The absence of heterogeneity in seroprevalence in this high transmission setting may have masked other potential relationships between exposure to enteric pathogens and distance to water. For example, among children 0 to 3 years old, the seroprevalence of *C. parvum* and *C. jejuni* were both very high (77% and 91% respectively). In a sensitivity analysis restricted to children younger than 12 months, there was an indication that the quantitative antibody levels for children living in the farthest quartile of distance compared to the nearest quartile of distance were higher for *V. cholerae* toxin beta subunit, *C. parvum* cp17 and cp23. However, the differences among this age sub-group were not statistically significant; the statistical power was likely limited by the lower number of children in this subset.

We were likely underpowered to determine differences in seroprevalence adjusted for socio-economic status. In the random 33% subset of children with available household asset information, children living in the furthest quartile of distance still had a higher seroprevalence of *S. enterica* and *G. intestinalis*, however the differences were not statistically significant.

There were several limitations of this study with respect to how the nearest water source was measured. First, we measured absolute linear distance rather than walking distance or time it takes to collect water. The study site region has tremendous gradation in altitude, with many high plateaus and steep valleys. In some cases, the distance to the nearest water source may not reflect the time it would take to ascend, descend or otherwise traverse the terrain. Second, we did not ask household which water source they were using. Households may use water sources that are further away via linear distance because of taste preference, ease of access, water source type or other reasons, namely terrain [33]. Third, the study site region is arid and there is variation in water availability by season. We simply measured the distance to the nearest water source at the time of the census and this may have not reflected a water source that was flowing and available at different times of the year. All of the above scenarios may have introduced non-differential misclassification of the exposure, which could bias associations towards the null. Finally, we opted to measure distance to the nearest protected or unprotected water source to evaluate the effect of water quantity on enteropathogen and *C. trachomatis* transmission. To evaluate the effect of water quality on enteropathogen transmission, distance to the nearest protected water source may have been a more appropriate exposure.

Another limitation of this study was the difficulty in determining seropositivity cut-offs for several of the antigens. The enteropathogens in particular pose a challenge. We were unable to determine reasonable cutoffs for *C. parvum* and *V. cholerae* using mixture models and had to discard *V. cholerae* from the seroprevalence analysis without a corresponding external negative control cutoff. Analyzing qualitative antibody levels is an alternative to seroprevalence that may retain the higher resolution needed in high-transmission settings [9]. When we evaluated differences in quantitative antibody levels according to distance to the nearest water source, the results were consistent with the seroprevalence findings for *S. enterica* LPS group B; quantitative antibody levels were also higher for ETEC and *G. intestinalis* but the differences were not statistically significant.

In conclusion, in this large population-based study of young children in the Amhara region of Ethiopia we document high transmission of *C. trachomatis, G. intestinalis, C. parvum, E. histolytica, S. enterica, C. jejuni* and ETEC. Children living furthest from a water source had higher seroprevalence of *S. enterica*, ETEC and *G. intestinalis* compared to children living closest to a water source. Serology was a useful approach to measure exposure to *C. trachomatis* and multiple enteropathogens. Our findings indicate the improving water quantity, through minimizing the distance to water collection, may reduce enteric pathogen transmission in settings such as Amhara with extreme water scarcity.

## Data Availability

Associated data will be available on OSF upon publication

## SUPPORTING INFORMATION

**Supplemental Figure 1:**
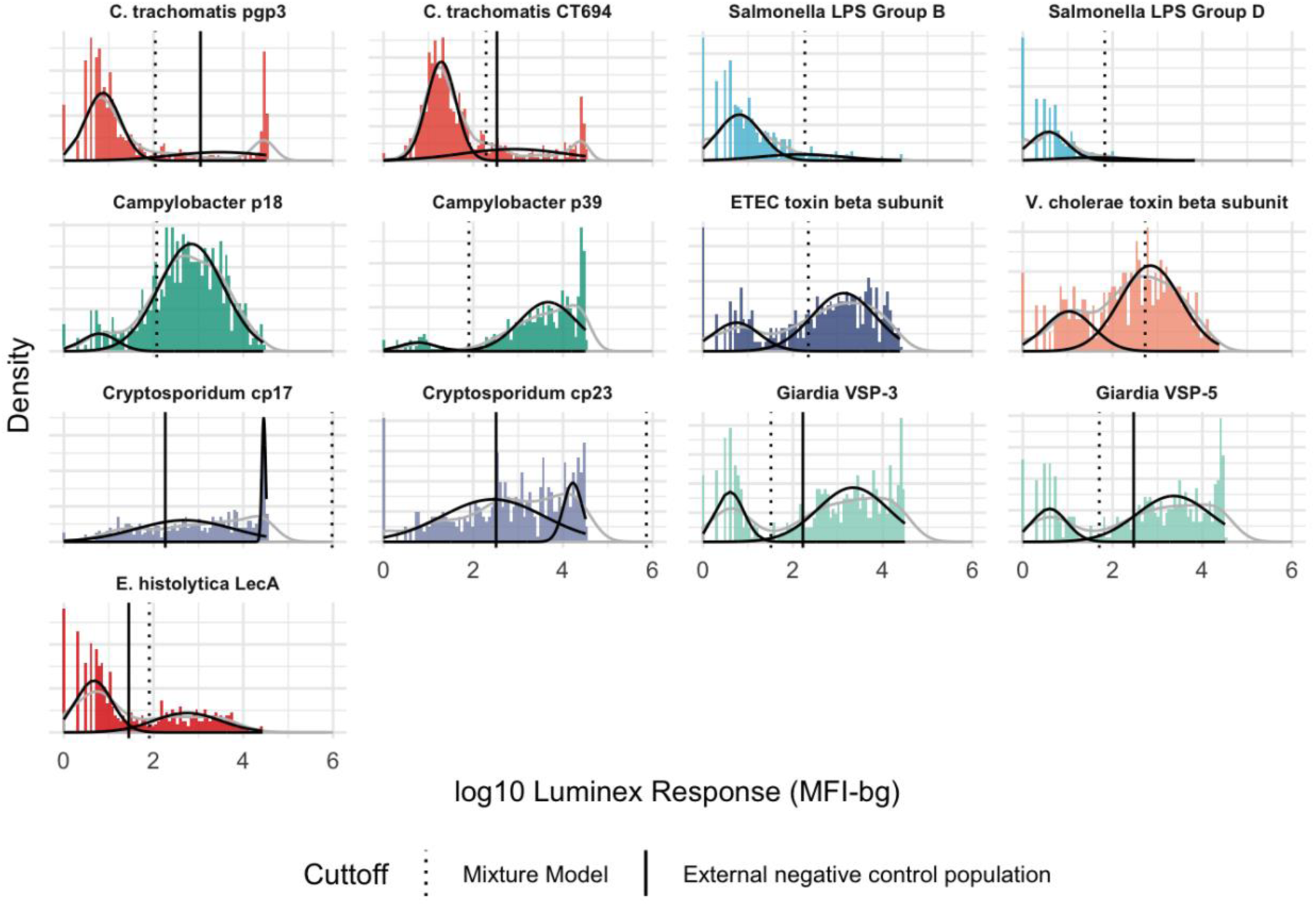
Distribution of IgG antibody response among children <24 months with ROC and mixture model cutoffs. IgG antibody response measured in multiplex using median fluorescence units minus background (MFI-bg) on the Luminex platform. Population restricted to children <24 months to derive cutoffs (n=317). Vertical lines mark seropositivity cutoffs based on external negative controls (solid) and finite Gaussian mixture models (dash). For Chlamydia trachomatis pgp3 & CT694 cutoffs were derived using recever operating characteristic (ROC) curves, for Cryptosporidium parvum Cp17 & Cp23 cutoffs were derived using a standard curve and for Giardia intestinalis VSP-3 & VSP-5 and Entomoeba histolytica LecA cutoffs were derived using the mean plus 3 standard deviations above a negative control panel.

**Supplemental Figure 2:**
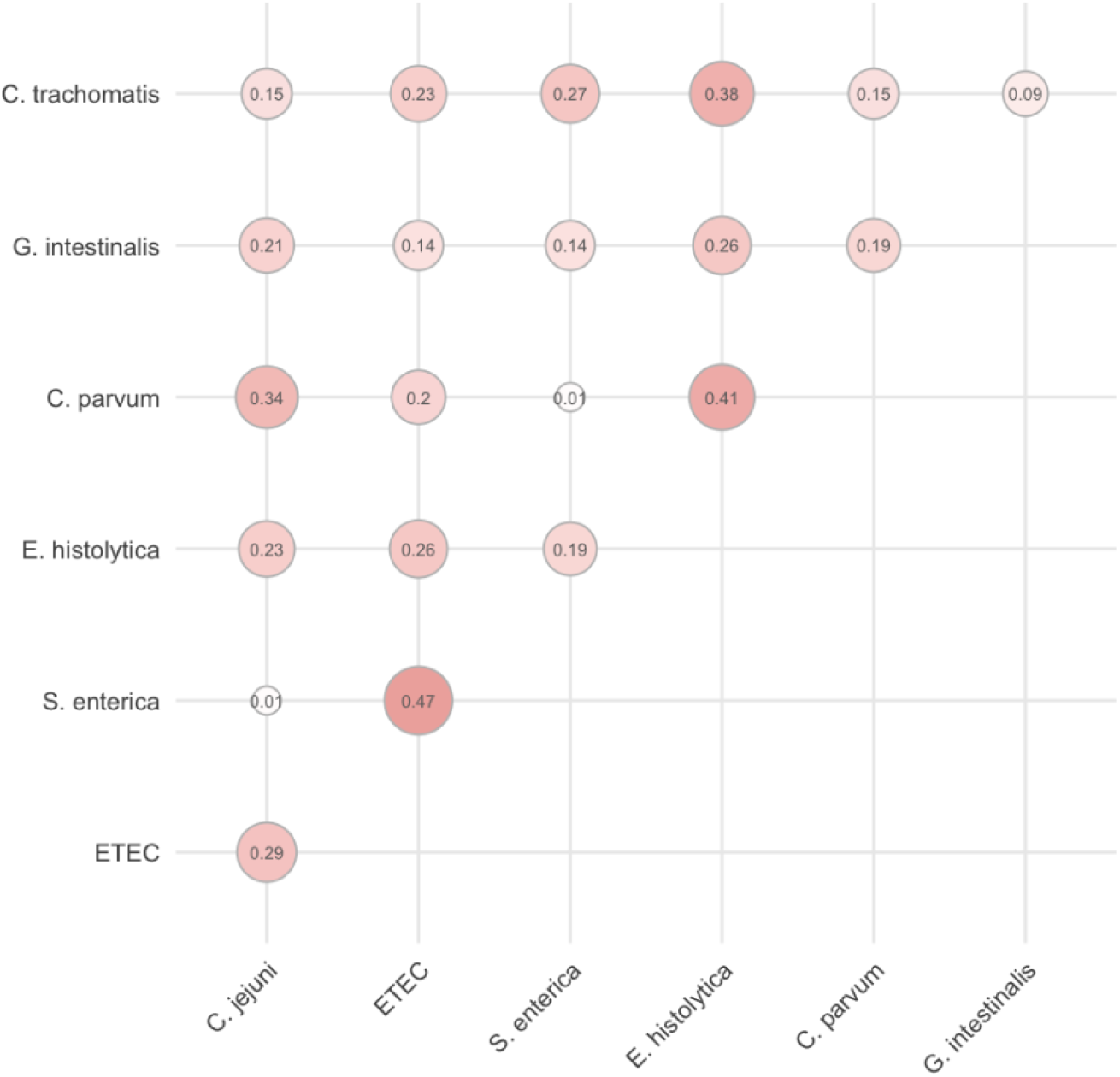
Community-level correlation in seroprevalence. Correlation between the mean community seroprevalence depicted with circles, greater circle area represents higher correlation. For pathogens with more than one antigen, positivity to antigen was considered positive. IgG response measured in multiplex using median fluorescence units minus background (MFI-bg) on the Luminex platform on 2328 blood samples from 2328 children aged 0 to 9 years.

**Supplemental Table 1:**
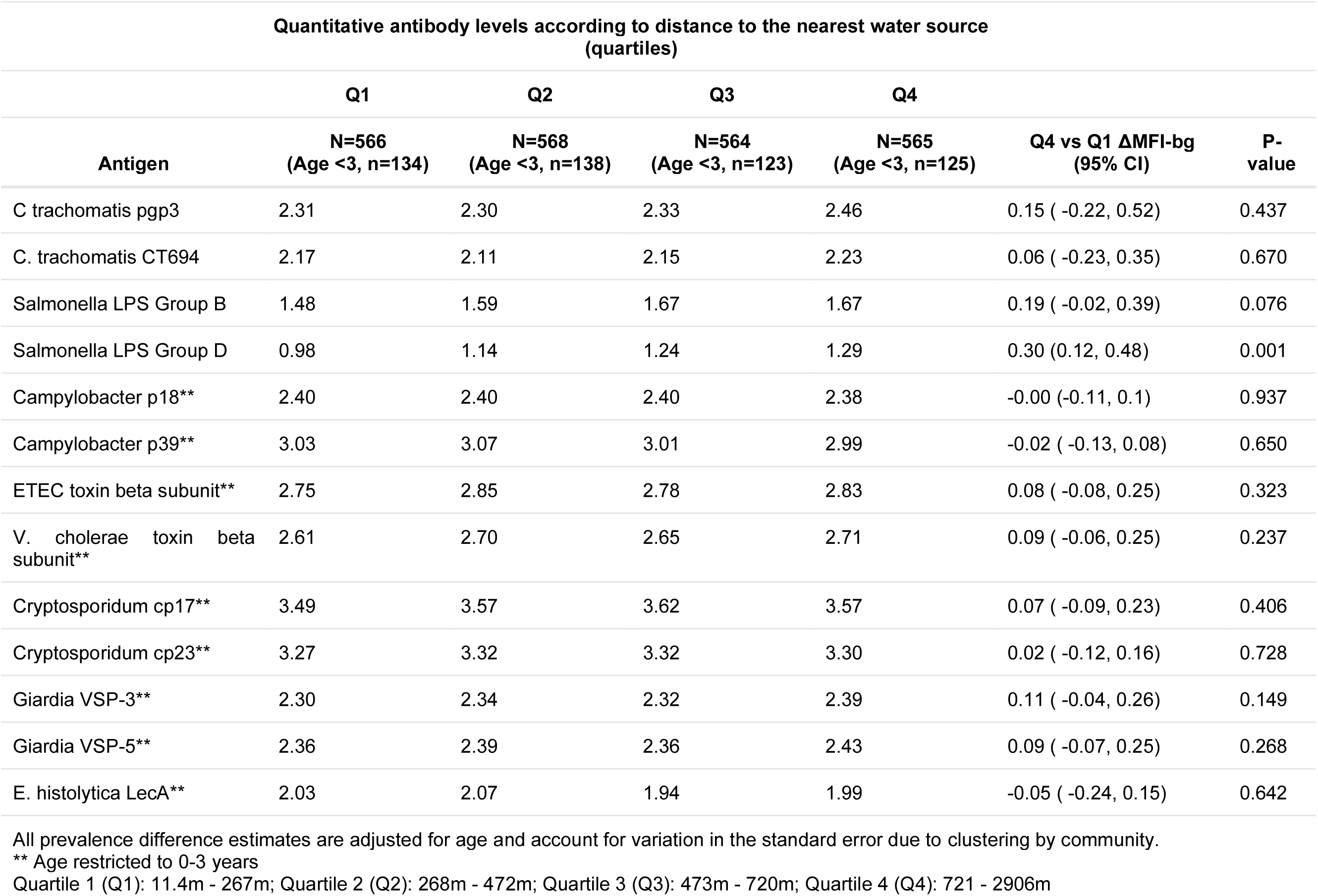
Quantitative antibody levels by distance quartile and differences comparing Quartile 4 to Quartile 1

| Quantitative antibody levels according to distance to the nearest water source (quartiles) |  |  |  |  |  |  |
| --- | --- | --- | --- | --- | --- | --- |
|  | Q1 | Q2 | Q3 | Q4 |  |  |
| Antigen | N=566<br>(Age <3, n=134) | N=568<br>(Age <3, n=138) | N=564<br>(Age <3, n=123) | N=565<br>(Age <3, n=125) | Q4 vs Q1 ΔMFI-bg<br>(95% CI) | P-value |
| C trachomatis pgp3 | 2.31 | 2.30 | 2.33 | 2.46 | 0.15 ( -0.22, 0.52) | 0.437 |
| C. trachomatis CT694 | 2.17 | 2.11 | 2.15 | 2.23 | 0.06 ( -0.23, 0.35) | 0.670 |
| Salmonella LPS Group B | 1.48 | 1.59 | 1.67 | 1.67 | 0.19 ( -0.02, 0.39) | 0.076 |
| Salmonella LPS Group D | 0.98 | 1.14 | 1.24 | 1.29 | 0.30 (0.12, 0.48) | 0.001 |
| Campylobacter p18** | 2.40 | 2.40 | 2.40 | 2.38 | -0.00 (-0.11, 0.1) | 0.937 |
| Campylobacter p39** | 3.03 | 3.07 | 3.01 | 2.99 | -0.02 ( -0.13, 0.08) | 0.650 |
| ETEC toxin beta subunit** | 2.75 | 2.85 | 2.78 | 2.83 | 0.08 ( -0.08, 0.25) | 0.323 |
| V. cholerae toxin beta subunit** | 2.61 | 2.70 | 2.65 | 2.71 | 0.09 ( -0.06, 0.25) | 0.237 |
| Cryptosporidium cp17** | 3.49 | 3.57 | 3.62 | 3.57 | 0.07 ( -0.09, 0.23) | 0.406 |
| Cryptosporidium cp23** | 3.27 | 3.32 | 3.32 | 3.30 | 0.02 ( -0.12, 0.16) | 0.728 |
| Giardia VSP-3** | 2.30 | 2.34 | 2.32 | 2.39 | 0.11 ( -0.04, 0.26) | 0.149 |
| Giardia VSP-5** | 2.36 | 2.39 | 2.36 | 2.43 | 0.09 ( -0.07, 0.25) | 0.268 |
| E. histolytica LecA** | 2.03 | 2.07 | 1.94 | 1.99 | -0.05 ( -0.24, 0.15) | 0.642 |

All prevalence difference estimates are adjusted for age and account for variation in the standard error due to clustering by community.
\*\* Age restricted to 0-3 years
Quartile 1 (Q1): 11.4m - 267m; Quartile 2 (Q2): 268m - 472m; Quartile 3 (Q3): 473m - 720m; Quartile 4 (Q4): 721 - 2906m

